# Predicted Norovirus Resurgence in 2021-2022 Due to the Relaxation of Nonpharmaceutical Interventions Associated with COVID-19 Restrictions in England: A Mathematical Modelling Study

**DOI:** 10.1101/2021.07.09.21260277

**Authors:** Kathleen M O’Reilly, Frank Sandman, David Allen, Christopher I Jarvis, Amy Gimma, Amy Douglas, Lesley Larkin, Kerry LM Wong, Marc Baguelin, Ralph S Baric, Lisa C Lindesmith, Richard A Goldstein, Judith Breuer, W John Edmunds

## Abstract

**Background:** To reduce the coronavirus disease burden in England, along with many other countries, the Government implemented a package of non-pharmaceutical interventions (NPIs) that have also impacted other transmissible infectious diseases such as norovirus. It is unclear what future norovirus disease incidence is likely to look like upon lifting these restrictions.

**Methods:** Here we use a mathematical model of norovirus fitted to community incidence data in England to project forward expected incidence based on contact surveys that have been collected throughout 2020-2021.

**Results:** We report that susceptibility to norovirus infection has likely increased between March 2020 to mid-2021. Depending upon assumptions of future contact patterns incidence of norovirus that is similar to pre-pandemic levels or an increase beyond what has been previously reported is likely to occur once restrictions are lifted. Should adult contact patterns return to 80% of pre-pandemic levels the incidence of norovirus will be similar to previous years. If contact patterns return to pre-pandemic levels there is a potential for the expected annual incidence to be up to 2-fold larger than in a typical year. The age-specific incidence is similar across all ages.

**Conclusions:** Continued national surveillance for endemic diseases such as norovirus will be essential after NPIs are lifted to allow healthcare services to adequately prepare for a potential increase in cases and hospital pressures beyond what is typically experienced.

## Background

In late January 2020 COVID-19 was first detected in the United Kingdom (UK) [1], and in response to increasing incidence and hospitalisations, the UK entered the first of three national lockdowns on the 23^rd^ of March 2020. The lockdowns, and other social restrictions resulted in a reduction in contact patterns in the community, as shown through self-reported contact rates collected using online surveys [2] and other metrics [3]. Subsequently, reports to national surveillance for many endemic diseases, including norovirus, declined across England throughout the rest of 2020 [4]. This reduction in reports of infectious disease has been consistent across many pathogens and observed in other countries where non-pharmaceutical interventions (NPIs) for the control of COVID-19 have been implemented (eg. Australia [5, 6] and the USA [7, 8]). With the introduction of vaccination against COVID-19 in England a considerable reduction in COVID-19 hospitalisations and deaths have been observed and this intervention has superseded the blunt tool of NPIs. The UK roadmap out of lockdown details the gradual relaxation of NPIs in 2021 [9], which will likely result in contact patterns more consistent with those seen prior to 2020.

Norovirus is an endemic viral infection present globally across high- and low-income countries with – as yet - no commercially available vaccine [10]. Norovirus infection can be asymptomatic or can result in gastrointestinal disease symptoms such as stomach cramps, diarrhoea and vomiting. In more vulnerable groups, such as the elderly or immunocompromised, norovirus can result in more severe and/or prolonged disease, and affected individuals may require hospitalisation for treatment. A recent analysis of hospital data in England ascribes an average of 40,800 (with an interquartile range (IQR) of 40,500-41,400) hospital episodes where norovirus was the primary diagnosis and a further 61,500 (IQR 58,700-62,500) with norovirus as a secondary diagnosis, meaning that norovirus is a significant public health and economic burden in England [11] and during hospital outbreaks can result in ward closures. Norovirus associated mortality within Europe has been estimated as 0.2 (95% uncertainty interval 0.1-0.2) deaths per 100,000 persons [12], which is at least 100-fold less than equivalent estimates for all-cause influenza illness [13]. Norovirus is the most common cause of gastrointestinal infections globally and has the highest burden of disease for intestinal infections in the UK [14]. Infections and outbreaks occur more frequently in the winter months, and in England are monitored through multiple national surveillance systems (eg. [15]) to ensure that unusual activity is detected, and that alerts to local health authorities are made for preventive actions. The probability of symptomatic disease is also influenced by host genetic factors [16], the infectious dose of norovirus [17], and previous exposure to infection. Norovirus genetic diversity is characterised by classifying isolates into genogroups, genotypes and strains (also termed variants) [18]; since 2012 the GII.4 genotype 2012 strain (shortened to “GII.4/2012”) has dominated in reports of symptomatic disease in England and globally. Prior to the GII.4/2012 strain, population strain replacement occurred every few years and was consistent across wide geographies. During these strain replacement events increased norovirus activity in the first year is typically observed, resulting in increased strain on health services [19]. While each dominant strain is termed dominant because it is isolated from a majority of cases in most settings, there is considerable genetic diversity of the remaining isolates, and the evolutionally impact of this remains unclear. The majority of norovirus infection is considered to occur via community transmission, and consequently changes in community contact patterns will likely influence norovirus transmission. However, norovirus has multiple routes of transmission; for example foodborne transmission is thought to contribute to approximately 16-35% in the UK, based on microbial risk assessments and detailed case investigations [20].

Here we illustrate through modelling the impact of COVID-19 NPIs used throughout 2020-2021 on the dynamics and incidence of norovirus disease in the community, leading on from a recent report of a reduction in norovirus activity [4]. We provide indicators of surveillance to inform what early warning of a norovirus resurgence may look like.

## Methods

### Modelling norovirus infection prior to the COVID-19 pandemic (up until March 2020)

We built an age-structured susceptible-exposed-infectious-recovered (SEIR) -like model that follows a previously developed model for norovirus, focussing on infections with the GII.4/2012 strain [21] (Figure 1). We assume a population of 100,000 individuals with age-structure similar to that reported in England, and that 20% of the population remain resistant to norovirus infection due to nonsecretor status of human histoblood group antigen carbohydrates [22]. Upon infection individuals are assumed to enter a short stage of exposure (or pro-dromal infection), and then become fully infectious and symptomatic for on average of 2 days [23, 24] (full details of the model, including differential equations and parameters, are given in “Additional file 1”). Individuals then enter the asymptomatic stage, where they remain moderately infectious [25] for on average 15 days when compared to the symptomatic stage but have no symptoms of disease [26]. Asymptomatic infection is assumed to correspond with norovirus shedding in stool that can be detected using RT-PCR. Upon recovery, individuals are immune to further symptomatic infection, but can develop asymptomatic infection. After an average of 5.1 years individuals are assumed to return to the susceptible class where infection will be symptomatic again [21]. To capture heterogeneity in transmission between ages, the transmission model is age-structured using eight age groups (0-4,5-14,15-24,25-34,35-44,45-54,55-64,65+). Symmetric contact rates between and within age groups prior to March 2020 were obtained for England from the European Polymod Study [27] and adapted for the age ranges used in here (using the *R* package *socialmixr*). All modelling and analyses were done in the software *R* (v 4.0.4).

**Figure 1.**
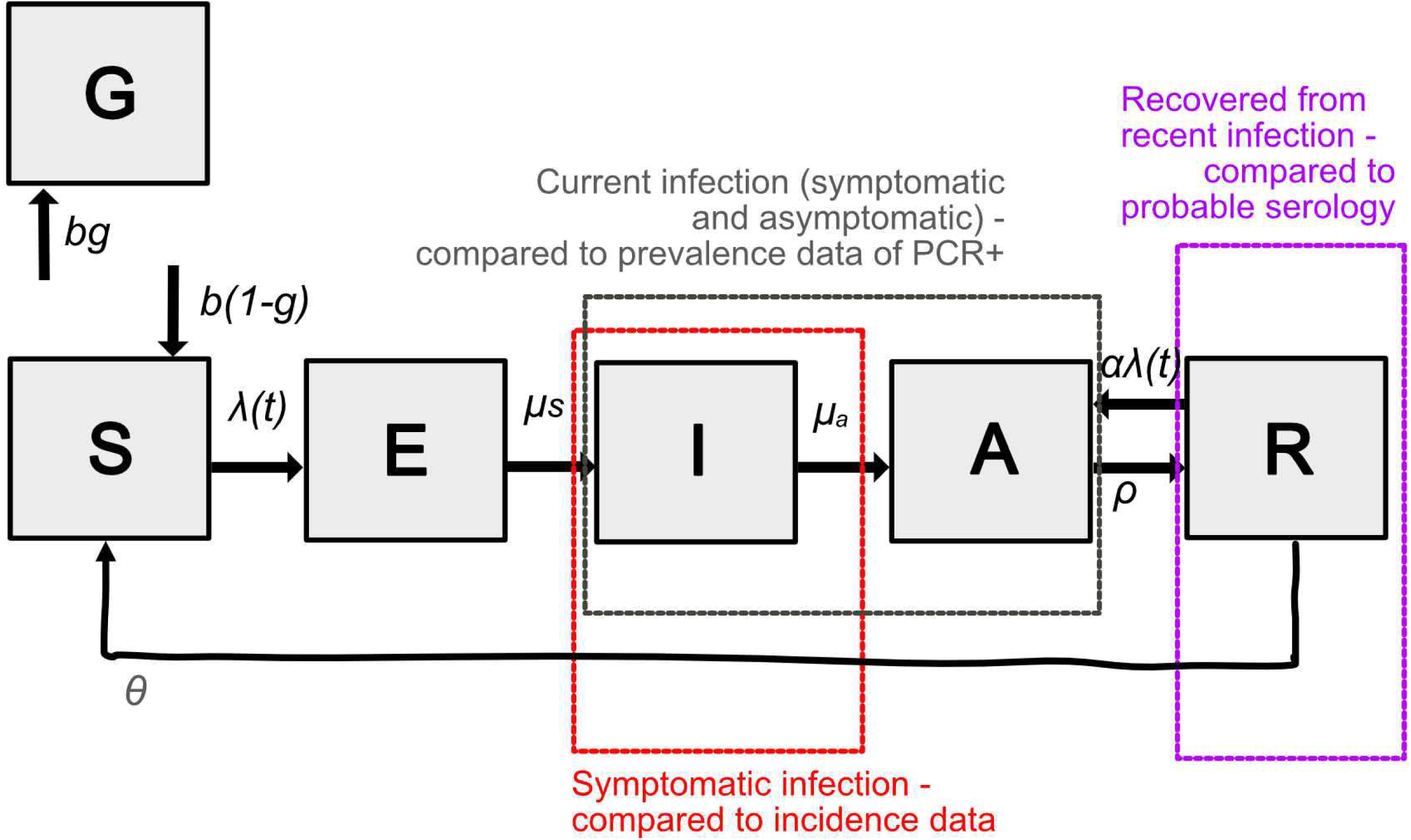
Schematic of the model for norovirus transmission. Each box represents a compartment of the system of equations; S – susceptible, E – pre-infectious, I – infectious and symptomatic, A – infectious and asymptomatic, R – recovered, G – genetically resistant to infection. Dashed boxes illustrate the assumed relationships between the model outputs and available data. The parameters specified in the figure are those used in the main model, see the supplementary information for additional assumptions tested as part of the sensitivity analysis.

A challenge of norovirus modelling is that there are broad observations in the epidemiology of norovirus which a model should ideally replicate, but these observations are from a wide spectrum of sources, with limitations and biases within each dataset. In the UK there have been two major studies in recent decades into infectious intestinal diseases (IID), which provide valuable information on community incidence. The first Study of Infectious Intestinal Disease (IID1) in England illustrated that norovirus is a common cause of IID in the community with considerable under-reporting to primary care [23]. A follow-on study (IID2) explored community incidence of gastrointestinal diseases (including norovirus), where incidence was highest in children and declined with age [28], and there was substantial under-reporting within the passive surveillance at the time. Serological data for norovirus is limited (due to challenges in establishing reliable correlates of protection [30] and enzyme immunoassays often being insensitive to strain-specific immunity), but where available, serological responses indicate seroconversion after acute illness [31] and suggest an increasing titre with exposure. In order to capture these general observations while needing to obtain reliable transmission rates within the community, we fit the probability of transmission given a contact to community age-stratified incidence data from IID2 [32] under different assumptions stated above, and also include a 20% under-reporting of symptomatic cases to account for a possible increase in incidence from the GII.4/2012 strain which has dominated symptomatic illness since this time. This one parameter is fitted using a Metropolis-Hastings algorithm, full details of the fitting procedure are provided in “Additional file 2”. For each scenario we provide estimates of incidence (ie. new cases of symptomatic infection) which were compared directly to incidence data. Additional model outputs were provided as a form of validation against what we would expect a realistic model of norovirus to output; estimates asymptomatic prevalence, a proxy for seroprevalence (assumed here to be the proportion of individuals that have recently recovered from symptomatic infection) and estimates of R_0_. Estimates of R_0_ were obtained for the pre-COVID-19 scenarios by running the model to endemic equilibrium and estimating R_0_ assuming R_0_ = 1/Pr(S). While the fit of the model to the data is the main criteria for model selection, we consider these additional metrics when making decisions on the appropriateness of each set of model assumptions.

While the above model assumptions align well with much of the evidence, there are uncertainties with some assumptions, and alternative assumptions may affect disease dynamics. As part of sensitivity analysis, we additionally include other models which capture the main uncertainties and compare the fit of these models to the data and other summary metrics (see “Additional file 3” for full details).

### Modelling norovirus infection during 2020-2021

From 23^rd^ March 2020 onwards we assume that the age-specific contact rates change as reported in a longitudinal cross-sectional study (‘Comix’) of in-person contacts in England [33]. Weekly measures of age-specific contact rates were collected from 24^th^ March 2020 and the study will continue to collect additional surveys until September 2021. The weekly contact rates are grouped into nine distinct periods of time including 3 different lockdowns that saw different degrees of NPIs being introduced and restrictions easing again with varying contact rates [34]. The model is first run to endemic equilibrium using pre-pandemic values of age-specific contact rates. Between 23^rd^ March 2020 until 19^th^ July 2021 a contact matrix from Comix is used for each period of time (specified in “Additional file 4”). For later projections we assume that contact rates return to pre-2020 values, an additional scenario that assumes adults only return to 80% of their pre-pandemic contact rates. We report the expected incidence of symptomatic norovirus infections in time, the proportion of the population susceptible to symptomatic infection. R_0_ was also estimated for each time-period; to do this we assume that only the contact rates have varied and so use the proportional change in the dominant eigenvalue of contact matrices (*ϑ*) to estimate *R*_*0*_(t), where R_0_(*t*) = *R*_*o*_/*ϑ*.

To support planning and national surveillance, we translate the incidence reported within the model to expected norovirus cases reported to the national Second Generation Surveillance System (SGSS). The SGSS is the national laboratory reporting system which records positive records of causative agents such as norovirus in England from frontline diagnostic laboratories [15]. As norovirus is not a notifiable causative agent in England under the Health Protection (Notification) regulations 2010 [35]. Data were extracted on the 28^th^ June 2021 for all specimen types from 1^st^ January 2014 onwards, and were deduplicated, providing an estimate of the weekly number of norovirus cases reported to SGSS. From the model the weekly incidence of symptomatic norovirus per 100,000 was scaled up to national incidence (using Office for National Statistics data on population size for England in 2019 of 56,290,000 individuals). Using the estimates within Tam et al. [14] we assume that for 287.6 (95% CI 239.1 to 346) symptomatic cases estimated nationally one case is reported within the SGSS. The data were compared to the model and the percentage difference reported, noting that the under-ascertainment reported in Tam et al. are from a laboratory reporting surveillance system in place prior to SGSS meaning that some changes in under-ascertainment are expected.

## Results

The age-specific incidence of norovirus observed in England is best replicated assuming that primary infection is symptomatic, the duration of asymptomatic infectiousness is 15 to 20 days, and that asymptomatic infection is considerably less infectious than symptomatic infection (Figure 2A and Table 1). Simulations that correspond to these observations resulted in a shedding prevalence less than 1%, the proportion of the population recently recovered from infection ranging from 24.54-30.55% and an R_0_ between 1.81-2.01. The age-specific incidence of symptomatic infection is highest in children aged <5 years, and incidence declines with age. When we compare this modelled incidence to the weekly SGSS five-year average (from norovirus year 2014/15 to 2018/19), a further 27% reduction in reporting is needed for cases to be equivalent (Figure 1B). Other models with different assumptions had a poorer fit to the data and were not taken further in the analysis.

**Figure 2.**
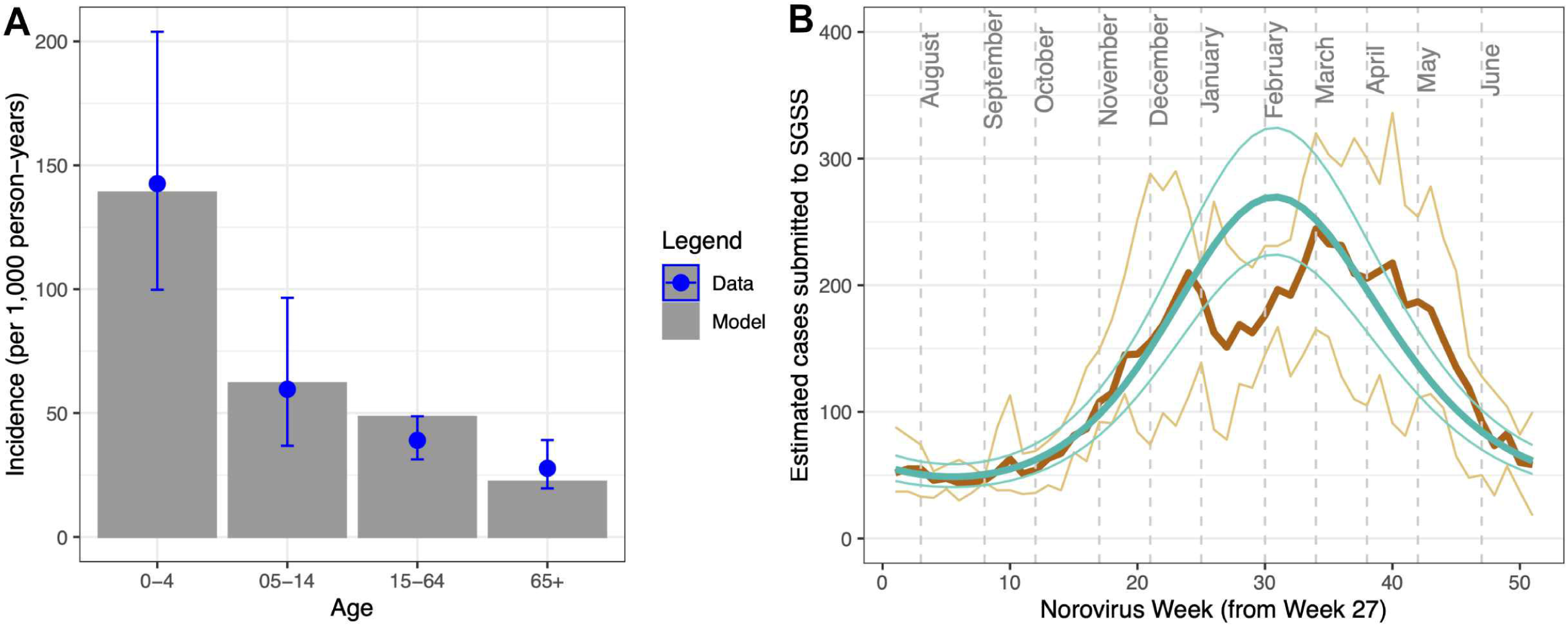
**Comparison of the norovirus model to A) data from Harris et al. of age-specific incidence of symptomatic norovirus infection where this fit was used to estimate the probability of transmission given a contact, B) weekly reported cases of norovirus reported to the SGSS system (thick brown line – 5 year average, thin brown lines – minimum and maximum).** The model incidence (per 100,000 person-years) was extrapolated to a national level, accounting for known under-reporting and under-ascertainment inherent in the surveillance data (287.6 (95%CI 239.1-346.0)) and a further 27% reduction in incidence in the model for alignment with the reported data. Dashed lines indicate the first day of each calendar month. SGSS: Second Generation Surveillance System.

**Table 1.** Summary of model assumptions trialled for norovirus, where each model was fitted to age-specific incidence data in England. Models A0-B20 were taken forward to estimate incidence of norovirus between 2020-2022.

| Model | A0 | A20 | B0 | B20 | D0 | D20 | E0 | F0 |
| --- | --- | --- | --- | --- | --- | --- | --- | --- |
| **Assumptions** |  |  |  |  |  |  |  |  |
| *First infection* | Always symptomatic | Always symptomatic | Always symptomatic | Always symptomatic | 50% asymptomatic | 50% asymptomatic | 50% asymptomatic | Always symptomatic |
| *Duration of Asymptomatic infectiousness (days)* | 15 | 15 | 20 | 20 | 15 | 15 | 15 | 15 |
| *Asymptomatic infectiousness relative to symptomatic* | 0.05 | 0.05 | 0.05 | 0.05 | 0.05 | 0.05 | 0.5 | 1 |
| *Under-reporting assumed in fitted data (%)* | 0 | 20 | 0 | 20 | 0 | 20 | 0 | 0 |
| **Results** |  |  |  |  |  |  |  |  |
| *Probability of transmission given a contact ($q_s$)* | 0.1892696 | 0.2070616 | 0.1693301 | 0.1824529 | 0.2183579 | 0.2193598 | 0.374627 | 0.1909186 |
| *Log-likelihood of age-specific incidence when fitted to data from England [15]* | -77.94 | -190.03 | -72.10 | -107.46 | -667.77 | -850.36 | -1458.08 | -1454.57 |
| *R0 (at endemic equilibrium)* | 1.81 | 2.03 | 1.81 | 2.01 | 1.94 | 1.62 | >10 | >10 |
| *Shedding prevalence (at endemic equilibrium) (%)* | 0.18 | 0.25 | 0.303 | 0.41 | 0.718 | 0.70 | 59.44 | 59.77 |
| *Sero-prevalence (at endemic equilibrium) (%)* | 24.64 | 30.55 | 24.54 | 29.92 | 27.73 | 37.80 | 20.49 | 20.17 |
| *Comments* | Good fit; used for 2020-2022 projections | Good fit; used for 2020-2022 projections | Good fit; used for 2020-2022 projections | Good fit; used for 2020-2022 projections | Poor fit to data, incidence in younger ages was under-estimated and incidence in older ages was over-estimated | Poor fit to data, incidence in younger ages was under-estimated and incidence in older ages was over-estimated | Poor fit to the incidence data (high incidence in adults) and unrealistic R0, but better comparison with expected seroprevalence and shedding | Poor fit to the incidence data (high incidence in adults) and unrealistic R0, but better comparison with expected seroprevalence and shedding |

From 23^rd^ March 2020 (“lockdown 1”) until 4^th^ September 2020 (the “School reopening” stage) the rate of infection for norovirus is sufficiently low that new infections are rare (Figure 2A), and an increase in population susceptibility is observed (Figure 2B). This period corresponds with minimal cases reported to the laboratory surveillance SGSS system (Figure S1). This period corresponded with an R_0_ for norovirus ranging from 0.43-0.77, meaning that transmission could not be sustained. The period between 4^th^ September 2020 to 5^th^ January 2021 corresponds to a higher average number of contacts within the population (up to 7.79 in average number of (unweighted) contacts), an increase of incidence in the model, and outside of the second lockdown and the Christmas period R_0_ was above 1.00 (see “Additional file 4”). This increase in incidence was not observed in the SGSS system. The subsequent “lockdown 3” period corresponds to a reduction in contacts (from 6.61 to 3.47) and the rate of infection falls to low levels again, until schools are re-opened on the 9^th^ March 2021 (Figure 2A). By this time point the model estimates that the proportion of the population susceptible to symptomatic infection has risen from 54% to 59% (a 9% increase) as a result of waning immunity (Figure 2B, age-specific values given in Table S1). Subsequently, model scenarios predict a rise in the rate of infection and a resurgence of norovirus in the community resulting in an annual incidence of cases up to 2-fold higher than simulations prior to 2020 (Figure 3A, full details in Table S1). This time period also corresponds with an R_0_ above 1.00, where the contact data collected while schools are corresponds to an estimated R_0_ of 1.28. However, this prediction is dependent upon assumptions on mixing patterns in the general population; if adult mixing is assumed to be 80% of pre-COVID levels an increase in incidence is not predicted. Instead, the rate of infection increases but at a lower level, resulting in a gradual increase in susceptibility in the population.

**Figure 3.**
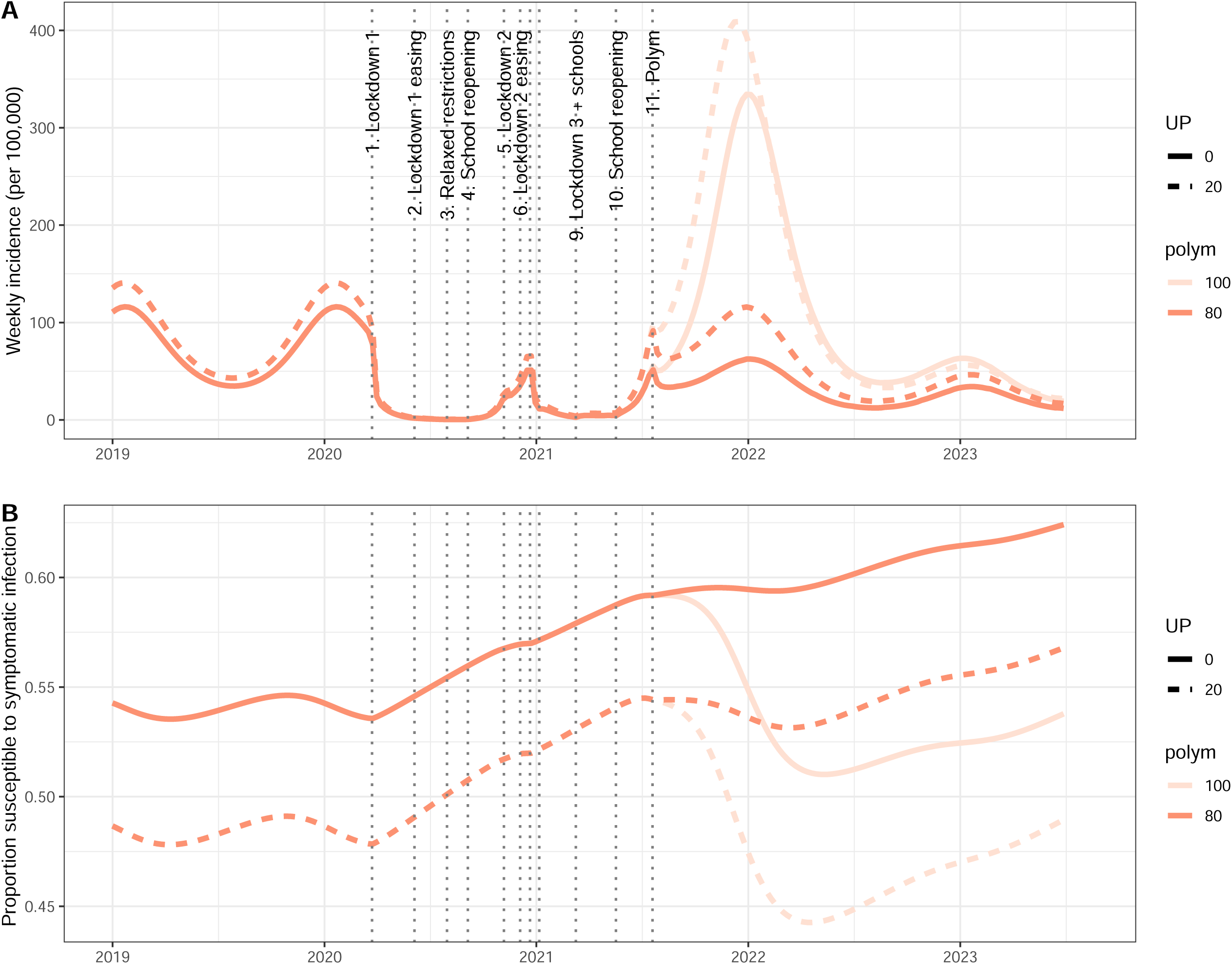
**Estimates of the impact of changing contact patterns due to COVID-19 restrictions on norovirus A) incidence and B) susceptibility to symptomatic infection from January 2019 to June 2023.** In each panel each colour represents simulations that assumptions contact patterns after July 2021 are the same as pre COVID-19 (light red) or adults have 80% fewer contacts (red), and allowing for different assumptions about under-reporting of norovirus incidence within Harris et al. [15]; solid lines assume no under-reporting and dashed lines assume 20% underreporting. UP: under-reporting, sim: simulated duration of asymptomatic infectiousness in days.

**Figure 4.**
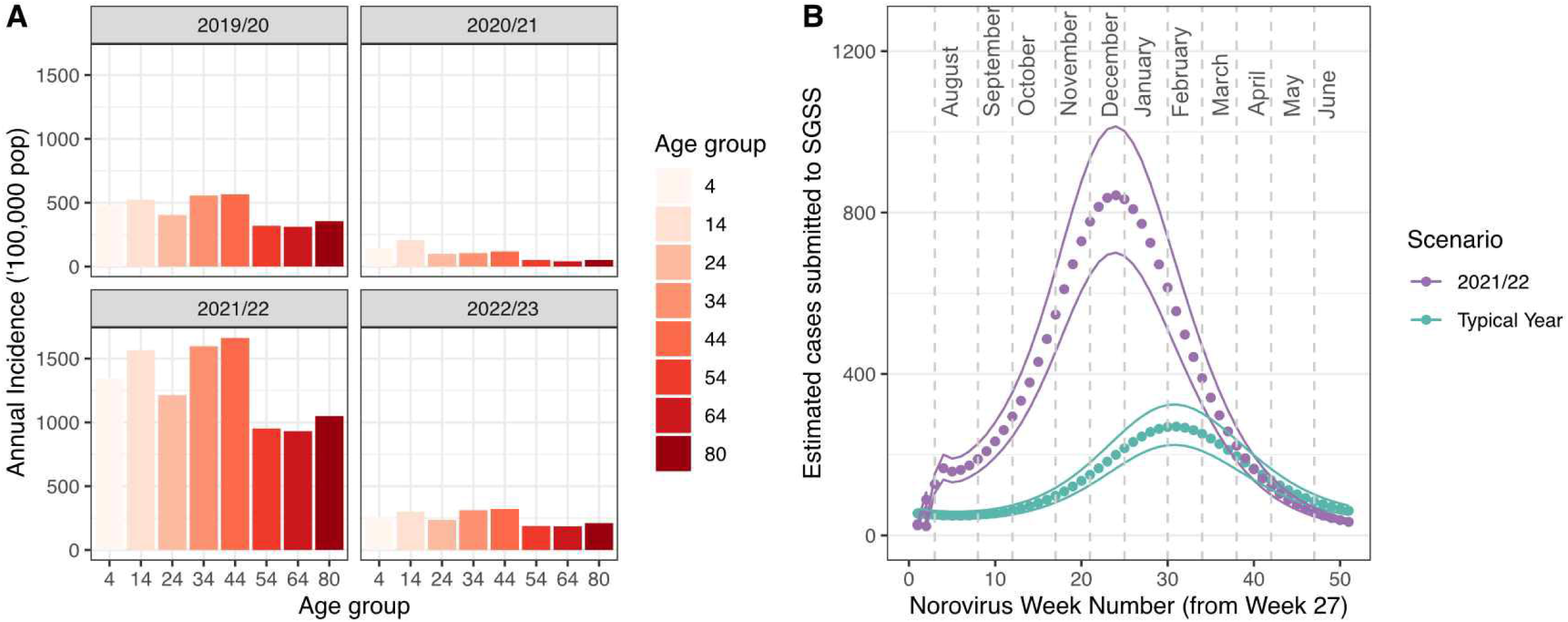
**Model estimates of A) incidence of symptomatic norovirus infections by age and norovirus year, B) predictions of cases reported within SGSS for the 2021/2022 year, compared with a typical norovirus year.** Simulations assuming a duration of shedding of 15 days and no under-reporting (outcomes from other simulations are shown in “Additional file 5”). Dashed lines indicate the first day of each calendar month. SGSS: Second Generation Surveillance System.

We also tested the impact of model assumptions regarding under-reporting and the duration of shedding (Figure S2). Assumptions about under-reporting affect the rate of infection; if incidence was under-reported in Harris [32] this will result in a higher rate of infection in the community and the potential for a more rapid change in incidence. Assumptions on the duration of shedding have a lesser impact; simulation assuming a shorter duration of shedding suggest a higher incidence in the 2021/22 norovirus year (Table S2).

## Discussion

The reduced reporting of norovirus cases and outbreaks that was observed in 2020 as a consequence of COVID-19-associated NPIs has been described within a number of countries, including England [4], Australia [5] and the USA [7, 8]. Our modelling study quantifies the potential unintended consequences of the build-up of individuals susceptible to norovirus infection in England, with epidemic potential for the autumn/winter of 2021 and 2022. Norovirus is an example of an endemic infectious disease where changes in contact rates can affect both the short and long-term incidence of symptomatic disease. This resurgence is likely to further increase “winter pressures” within the NHS that may lead to more hospital admissions due to norovirus, hospital outbreaks of norovirus with bed-days lost as beds are kept empty for infection prevention and control, and treatment delays for non-norovirus patients [11, 36], therefore preparations for mitigating the effect of an increase in incidence are essential. This is especially true at the current time where there has been considerable disruption to healthcare services due to the COVID-19 pandemic and there are a record number of 5 million people on waiting lists to receive hospital care following the impact of COVID-19 in England [37]. While there are several caveats of the modelling to consider in the interpretation of this analysis, this is the first analysis of disease dynamics for norovirus affected by changes in NPIs.

More broadly, the unintended consequences of NPIs are likely to be widespread across other endemic diseases (eg. respiratory syncytial virus and influenza) where incidence pre-COVID-19 was limited by populations being largely immune to infection [38, 39]. A challenge in interpretation of the likely future trajectory of incidence is quantification of how NPIs impact physical contacts in the community and how this impacts transmission of infectious disease. Here we make use of the longitudinal cross-sectional study of contacts, which has been instrumental in predicting incidence of COVID-19 [40], and show that these data can be very useful for quantifying dynamics of endemic diseases in addition to COVID-19. Continued collection of these data and incorporation into models of infectious disease will be essential in informing our understanding of disease transmission as the pandemic continues to unfold. It is likely that the impact of NPIs on norovirus, respiratory syncytial virus and influenza was more pronounced than for SARS-CoV-2 because the effective reproduction number (R_e_, equivalent to R_0_ multiplied by the proportion susceptible) for these endemic pathogens is close to 1.00. Theoretically, the NPIs are designed to limit contact rates, which will reduce R_0_ and in turn R_e_. At least in the period of time that has been experienced, this will limit transmission and disease incidence, as we have observed. The impact of further restrictions should still be effective, but with increases in susceptibility the indirect effect of NPIs on endemic diseases could become less effective with time Our analyses make the implicit assumption that contacts described within surveys correlate with contacts relevant for infectious disease transmission where the primary mode is direct contact; while this is apparent for infections such as influenza and SARS-CoV-2, there is less evidence for norovirus. These assumptions explain why the rate of infection is predicted to increase when contacts correspond to an R_0_ above 1.00. The reported incidence of norovirus in the coming months, the changing behaviours of the community in contacts and infection prevention, and the actions public health officials will take to prevent a large epidemic, will be the test of this hypothesis. As the COVID-19 pandemic evolves, potentially including more transmissible and vaccine-evading variants, NPIs that include the ongoing working-from-home recommendation through to school closures and lockdowns, remain possible. A major assumption of our model, which reflects current understanding of norovirus epidemiology, is that contacts with and between children under 5 years are the main driver of norovirus transmission. Consequently, any changes in contacts with this age group will have a large effect on norovirus dynamics.

In the analysis the incidence of norovirus reported was replicated well in the simulations, with the exception of the expected increase in cases at the end of 2020. This potentially suggests that the rate of infection assumed in the model is higher than is experienced in the community, despite the fact that the rate of infection was fitted to community data. Alternatively, the effect of reduced to the SGSS due to changes in healthcare seeking behaviours and reduced laboratory testing of samples obtained may have impacted on the comparator data. This observation means that some caution should be applied to the findings presented here as the model predictions are sensitive to assumptions about contact rates and the probability of transmission, which remain uncertain. Further work will explore this in more detail through the use of additional contact surveys and exploration of other datasets that can improve our understanding of norovirus transmission in the community.

The majority of norovirus disease occurs within the community; there is a significant disease burden resulting in school closures and days of work lost (especially as infection can be experienced relatively frequently) [42]. Hygiene measures, such as hand washing with soap, enhanced environmental cleaning and staying at home when ill, have been part of usual recommended practice to reduce norovirus transmission, but the COVID-19 pandemic has indirectly enforced the adherence to these measures. It is possible that beyond 2021 these measures may remain as usual practice in the short to medium term, which may well mean that incidence of infectious diseases such as norovirus is lowered compared with historical norms. An additional challenge of norovirus largely being a community illness is that norovirus surveillance is affected by both under-ascertainment and under-reporting, resulting in considerable uncertainty in true estimates of burden. With an increase in community transmission of norovirus, we would expect an increase in incidence, including severe disease in institutional settings (such as care homes for the elderly). However, much of the COVID-19 interventions targeted at institutional settings may also limit the incidence of norovirus; the impact of these interventions on norovirus warrants investigation. While we compared the model output to cases reported into the SGSS, surveillance is biased towards cases within hospitals, which will be a combination of community and hospital acquired infections. For this reason we have fitted to data from a community cohort study and used the SGSS data as a validation of the model estimates. The data from IID2 is over 10 years old, and may not fully represent incidence experienced in recent years, due to strain replacement in 2012 and hypothesised higher incidence of infection. Indeed, a recent community cohort study in the Netherlands report an incidence of symptomatic norovirus of 339.4 cases per 1000 person-years [43], which is higher than that reported in the IID2. We aimed to overcome the weaknesses in data by having multiple models with varying assumptions to cover a wide range of plausible present-day scenarios of norovirus dynamics. We identified a difference between model predictions and SGSS of 73%, which is well within a plausible range.

A further complexity in norovirus epidemiology is the possibility of strain replacement in the coming months or years; the GII.4/2012 strain has been in circulation in England since 2013 and could continue to dominate, but the emergence of other variants also have potential [44]. The likelihood and how a new variant may impact norovirus dynamics is currently uncertain and is likely to depend on the relative virulence of a new strain and the heterotypic immunity from infection with the GII.4/2012 strain. Further work will include consideration of strain-specific immunity profiles within the community to understand the impact of multi-strain dynamics and the potential impact of vaccination.

## Conclusions

This modelling study suggests that norovirus incidence in the community is likely to remain approximately similar to or substantially increase beyond what has been experienced in years prior to 2020. The results are sensitive to future contact patterns in the community, and the adherence to preventive actions, which will affect the probability of transmission. The lower incidence of norovirus reported in surveillance aligns with model assumptions on reduced contact rates from March 2020 to June 2021, and is consistent with an increase in population susceptibility. The short to long term impact of this increased susceptibility places populations at risk of norovirus disease, but the scale of the impact remains to be seen.

Continued investment to maintain robust national surveillance systems will remain critical to enable measures to limit the impact of these resurgences and provide essential information to public health bodies to support implementation of preventive actions.

## Data Availability

The code and data used to conduct these analyses are found at https://github.com/kath-o-reilly/norovirus_NPIs. For data access to the SGSS data used in the study applicants can apply to the PHE team by emailing

## Abbreviations

IID: Infectious Intestinal Disease
NPI: Non pharmaceutical interventions
SEIR: susceptible-exposed-infectious-recovered
SGSS: Second Generation Surveillance System
UK: United Kingdom

## Declarations

### Ethics approval and consent to participate

Institutional ethics approval was not sought because this is a retrospective study and the databases used are anonymised and free of personally identifiable information. Consent was not sought as the data are part of ongoing public health surveillance.

### Consent for publication

not applicable

### Authors’ contributions

KO, FS, JB, and JE conceived the study. KO, CI, AG, AD, LL, KW carried out data analysis and KO carried out the modelling. Interpretation of the data findings were done in discussion with KO, FS, DA, CJ, AD, LL, LL, RG JB and JE, and all authors contributed to the final draft of manuscript, and approved the for publication.

## Acknowledgements

The authors are grateful for discussions within the wider Noropatrol consortium. The authors also acknowledge and are grateful for the data obtainable from public health diagnostic laboratories in England that submit laboratory reports of norovirus into SGSS.

## Additional Files

**<u>Additional file 1 (.docx)</u>**

**Full description of the mathematical model for norovirus**

**<u>Additional file 2 (.docx)</u>**

**Details of ‘Polymod’ and ‘Comix’ contact matrices**

**<u>Additional file 3 (.docx)</u>**

**Alternative model assumptions for norovirus**

**<u>Additional file 4 (.docx)</u>**

**Details of ‘Polymod’ and ‘Comix’ contact matrices**

**<u>Figure S1 (.doc)</u>**

**Comparison of simulations and cases reported to SGSS between July 2019 to June 2021**

**<u>Figure S2 (.doc)</u>**

**Investigating the impact of reduced contact patterns from July 2021 compared to pre-pandemic**

**<u>Table S1 (.doc)</u>**

**Estimated annual age-specific norovirus incidence (2019-2022)**

