## Supplementary material for "Predicted Norovirus Resurgence in 2021-2022 Due to the Relaxation of Nonpharmaceutical Interventions Associated with COVID-19 Restrictions in England: A Mathematical Modelling Study": AF 1

**Additional file 1**

**Full description of the mathematical model for norovirus**

We built an age-structured SEIR-like model that follows a previously developed model for norovirus, focussing on GII.4 infections [21]. We assume a population of 100,000 individuals with age-structure similar to that reported in England, and that 20% of the population remain resistant to norovirus GII.4 infection due to nonsecretor status of human histoblood group antigen carbohydrates [22]. Births are assumed to either enter the genetically resistant class with probability g (G in Figure 1), or the susceptible class (S) with probability 1-g. Upon infection individuals are assumed to enter a short stage of exposure (or pro-dromal infection), and then become fully infectious and symptomatic for on average 2 days [23, 24]. The rate of infection is assumed to be a function of the proportion of individuals in the symptomatic and infectious class and the asymptomatic, where the relative contribution of the asymptomatic individuals is defined by $\delta$ [25]. Individuals then enter the asymptomatic stage, where they remain moderately infectious for on average 15 days when compared to the symptomatic stage but have no symptoms of disease [26]. Asymptomatic infection is assumed to correspond with norovirus shedding in stool that can be detected using PCR. Upon recovery, individuals are immune to further symptomatic infection, but can develop asymptomatic infection. After an average of 5.1 years individuals are assumed to return to the susceptible class where re-infection will be symptomatic again [21]. We make the simplistic assumption of there being one norovirus variant present in the population where immunity to further infection impacts only the probability of being symptomatic.

**Table S1. Parameters within the mathematical model for norovirus and sources of values used.**

| Symbol | Parameter | Values assumed here and source (brackets indicate source from main paper) |
| --- | --- | --- |
| ${\mathbf{1/}\boldsymbol{\mu}}_{\boldsymbol{s}}$ | Duration of incubation | 1 day |
| ${\mathbf{1/}\boldsymbol{\mu}}_{\boldsymbol{a}}$ | Duration of symptoms | 2 days [24, 25] |
| $\mathbf{1/}\boldsymbol{\rho}$ | Duration of asymptomatic virus shedding | 15 days [26] |
| $\boldsymbol{\delta}$ | Relativeness infectiousness during incubation and asymptomatic period | 0.05 [25] |
| $\mathbf{1/}\boldsymbol{\theta}$ | Duration of immunity | 5.1 years [21] |
| $\boldsymbol{\alpha}$ | Relative rate of infection for individuals who have recently recovered from symptomatic infection, compared to symptomatic infection | 1 |
| *b* | Birth rate | Tuned to maintain population size of 100,000* |
| *g* | Proportion of births assumed to be resistant to GII.4 infection | 0.2 [22] |
| *f* | Proportion of first infections that are assumed to be symptomatic | 1, but varied as part of sensitivity analysis |
| $\boldsymbol{\eta}$ | Age-specific death rate | Tuned to fit to ONS census data |
| $\boldsymbol{c}_{\boldsymbol{ij}}$ | Age-specific contact rates, where j is in contact with i | Polymod and Comix data, assuming symmetric contact rates |
| *q* | Probability of transmission per infectious contact | Estimated from data |

* But note that the impact of a reduced birth rate in 2020-2021 was also trialled in simulations
