## Supplementary material for "Predicted Norovirus Resurgence in 2021-2022 Due to the Relaxation of Nonpharmaceutical Interventions Associated with COVID-19 Restrictions in England: A Mathematical Modelling Study": AF 2

**Additional file 2**

**Description of the model fitting procedure**

The available data on age-specific incidence consists of 4 datapoints (age-specific incidence in units of 1000-person-years), consequently only the probability of transmission given an infected contact (*q*) is estimated from the data. We assume that the likelihood of the model output (age-specific incidence) given the data are normally distributed. The best fitting value of *q* is determined using a Metropolis-Hastings (MH) algorithm, using the following inputs;

- Priors for *q* are assumed to be log-normally distributed with mean of 0.18 and standard deviation 0.5, resulting in wide 95% CI intervals of 0.064-0.490.
- Proposed values for *q (q’)* are sampled from the current accepted value and a standard deviation of 0.003 (providing an acceptance rate varying from 0.5-0.7, depending upon simulations)
- Iterations of the MH algorithm were run for at least 1,000 iterations, where convergence was assessed by there being no meaningful reduction in the log-likelihood with additional iterations run.
- New values for *q* were accepted using probability $A\left( q^{'},q_{t} \right)=min\left( 1,\frac{P\left( q^{'} \right)g(q_{t}|q^{'})}{P\left( q_{t} \right)g(q^{'}|q_{t}} \right)$

For each of the models that represent different assumptions about norovirus natural history, a comparison of the log-likelihood was performed. Formally, the model with the largest log-likelihood would be the model to take forward to further simulations, and models with a difference in log-likelihood above 3.84 (p<0.05) should be excluded. However, several models (A0, B0 and A20, B20) have no biologically meaningful difference in reported incidence, and the additional outputs (seroprevalence, asymptomatic prevalence and R_0_) are not meaningfully different either, so we make the pragmatic decision to take all models forward to further simulations as we feel that the variability in projected incidence is an important consideration. In this circumstance the model structures are all ‘nested’, for example by setting a parameter to zero one model structure can become another model structure, meaning that this simple log-likelihood comparison between model fits is possible. In other circumstances where nesting is not feasible, other approaches would be required, such as computing Bayes factors of models (and identifying the Bayes factors using thermodynamic integration).
