## Supplementary material for "Predicted Norovirus Resurgence in 2021-2022 Due to the Relaxation of Nonpharmaceutical Interventions Associated with COVID-19 Restrictions in England: A Mathematical Modelling Study": AF 3

**Additional file 3**

**Alternative model assumptions for norovirus**

The model structure developed by Lopman et al. [19] was our starting point for a model of norovirus transmission. Among the research group discussions of the model assumptions led us to trial alternative hypotheses by varying the model structure and comparing the model fit to the data. The hypotheses we explored are stated below with details of the justification. The log-likelihood fit of the model to the data is used to explore the evidence for using this model in further simulations; if the fit is substantially worse the model is not taken further. However, it should be acknowledged that this is not an exhaustive exploration of these hypotheses; the available data only enables us to vary one parameter at a time. With additional evidence these hypotheses should be further explored.

*B. the duration of asymptomatic shedding is on average 20 days in duration*

Studies where individuals with recent norovirus infection or in controlled experiments indicate that norovirus can be shed up to 3 weeks after the onset of illness [17]. The duration of shedding varies between individuals, and the quantity of virus shed also varies throughout the time-course of infection.

*D. The first norovirus infection can be asymptomatic*

While asymptomatic infection is well documented in older ages, it is less evident in younger ages where one might be confident that the infection is the first experienced. Within Han et al [46] a norovirus outbreak is reported within a paediatric ward where asymptomatic infection is reported but the extent of infections are not specified. Reports from paediatric wards are very useful in understanding the natural history of first infections, but children within paediatric wards may not be representative of the general population due to the medical reasons for them being present on the ward. To test this hypothesis we specify a model where a proportion of births (*f*) are assumed to be ‘born into’ the recovered class, resulting in the first infection being asymptomatic.

*E. Asymptomatic infection is as infectious as symptomatic infection*

The symptoms of norovirus infection (vomiting and diarrhoea) likely facilitate transmission to susceptible individuals through direct and fomite contact, and outbreak investigation has illustrated that asymptomatic individuals do not meaningfully contribute to transmission [25]. The default model assumes that for any given time period, asymptomatic infection is 5% as infectious as symptomatic infection. However, in a community setting, especially where parents may be caring for infected children, the evidence for asymptomatic transmission is less clear. To capture this alternative hypothesis that especially in community settings transmission of asymptomatically infected people may have a considerable contribution to transmission, we trial alternative values of relative infectiousness ranging from 0.05 to 1.0, and assess the fit of the model to the data.

[46] Mi Seon Han, Seung Min Chung, Eun Jin Kim, Chan Jae Lee, Ki Wook Yun, Pyoeng Gyun Choe, Nam Joong Kim, Eun Hwa Choi. Successful control of norovirus outbreak in a pediatric ward with multi-bed rooms, American Journal of Infection Control, Volume 48, Issue 3, 2020, Pages 297-303, ISSN 0196-6553, https://doi.org/10.1016/j.ajic.2019.07.022.
