## Supplementary material for "Predicted Norovirus Resurgence in 2021-2022 Due to the Relaxation of Nonpharmaceutical Interventions Associated with COVID-19 Restrictions in England: A Mathematical Modelling Study": AF 4

**Additional file 4**

**Details of ‘Polymod’ and ‘Comix’ contact matrices**

The dates provided here are the corresponding dates used in Jarvis et al [2].

| **Date** | **Corresponding matrix** | **Average number of contacts (unweighted)** | **Estimate of R_0_*** |
| --- | --- | --- | --- |
| 01/01/2020 | 0. Polym | 11.12 | 1.81 |
| 23/03/2020 | 1. Lockdown 1 | 3.23 | 0.43 |
| 04/06/2020 | 2. Lockdown 1 easing | 3.96 | 0.55 |
| 30/07/2020 | 3. Relaxed restrictions | 5.5 | 0.77 |
| 04/09/2020 | 4. School reopening | 7.79 | 1.29 |
| 05/11/2020 | 5. Lockdown 2 | 5.87 | 0.99 |
| 03/12/2020 | 6. Lockdown 2 easing | 6.61 | 1.07 |
| 20/12/2020 | 7. Christmas | 3.51 | 0.47 |
| 05/01/2021 | 8. Lockdown 3 | 3.47 | 0.48 |
| 09/03/2021 | 9. Lockdown 3 + schools | 5.65 | 0.91 |
| 17/05/2021 | 10. School reopening | 7.79 | 1.28 |
| 19/07/2021 | 11. Polym (if 80% of contacts in adults) | 9.17 | 1.52 |
| 1/07/2023 | End of simulation |  |  |

* R_0_ is estimated for each period of time by finding the proportional difference in the dominant eigenvalue of the symmetrical contact matrix when compare to the “Polymod” contact matrix used prior to NPIs associated with the COVID-19 pandemic.
