## Supplementary material for "Predicted Norovirus Resurgence in 2021-2022 Due to the Relaxation of Nonpharmaceutical Interventions Associated with COVID-19 Restrictions in England: A Mathematical Modelling Study": S1

**Figure S1**

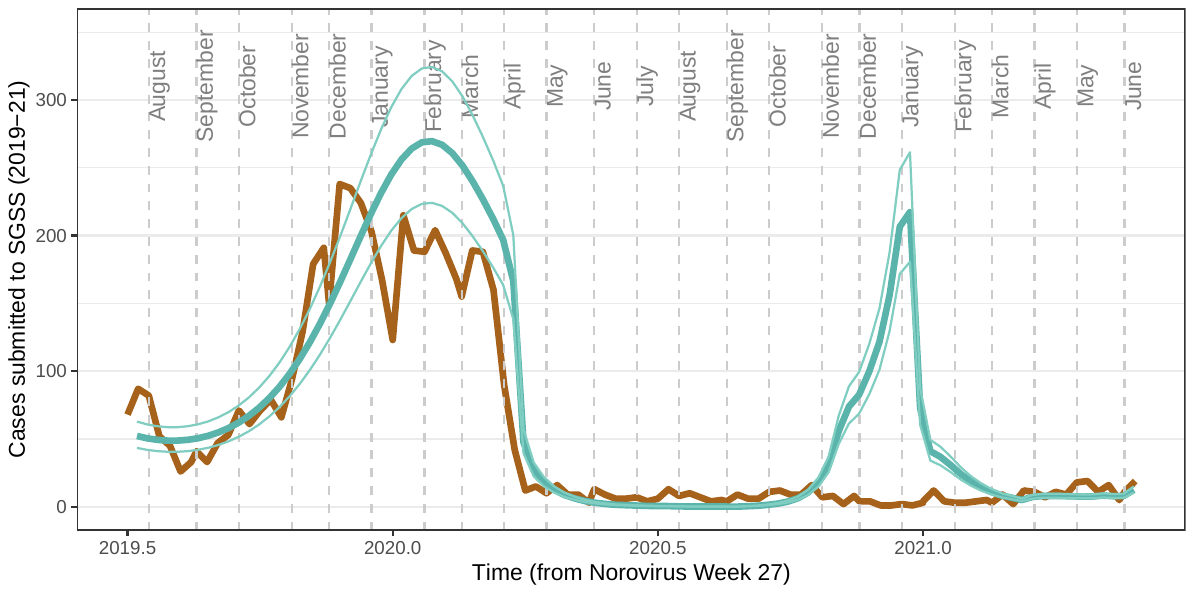

Figure S1. Comparison of simulations (green lines) and cases reported to SGSS (brown line) between July 2019 to June 2021.
