## Supplementary material for "Predicted Norovirus Resurgence in 2021-2022 Due to the Relaxation of Nonpharmaceutical Interventions Associated with COVID-19 Restrictions in England: A Mathematical Modelling Study": S2

**Figure S2**

**Investigating the impact of reduced contact patterns from July 2021 compared to pre-pandemic**

**Figure S2. Estimates of the impact of changing contact patterns due to COVID-19 restrictions on norovirus A) incidence and B) susceptibility to symptomatic infection from January 2019 to June 2023.** In each panel each colour represents simulations assuming a duration of asymptomatic infectiousness of 15 (light red) or 20 (red) days, and allowing for different assumptions about under-reporting of norovirus incidence within Harris et al. [15]; solid lines assume no under-reporting and dashed lines assume 20% underreporting. UP: under-reporting, sim: simulated duration of asymptomatic infectiousness in days.
